# Unique *SLC26A4* Mutation Spectrum in Mongolian Patients with Enlarged Vestibular Aqueduct: A Whole-Exome Sequencing Study

**DOI:** 10.1101/2025.02.03.25321220

**Authors:** Jargalkhuu Erdenechuluun, Bayasgalan Gombojav, Tserendulam Batsaikan, Yue-Sheng Lu, Narandalai Danshiitsoodol, Zaya Makhbal, Maralgoo Jargalmaa, Tuvshinbayar Jargalkhuu, Ho-Peng Hsu, Pei-Hsuan Lin, Hung-Ju Su, Chien-Hsing Lin, Chuan-Jen Hsu, Pei-Lung Chen, Cheng-Yu Tsai, Chen-Chi Wu

## Abstract

**Background:** Enlarged vestibular aqueduct (EVA) is a common inner ear malformation that can cause sensorineural hearing loss. It is frequently associated with mutations in the *SLC26A4* gene. This study aimed to investigate the genetic basis of hearing loss in Mongolian patients with EVA.

**Methods:** Whole exome sequencing was performed in 19 Mongolian patients from 15 unrelated families diagnosed with EVA with/without cochlear incomplete partition type II. All patients underwent high-resolution computed tomography of the temporal bone to confirm the diagnosis.

**Results:** Biallelic *SLC26A4* pathogenic variants were identified in all 15 families (100%). The most frequent variant was c.919-2A>G (40%), followed by c.2027T>A (23.3%) and c.1318A>T (16.7%). Population-specific variants in East Asians (c.919-2A>G), North Asians (c.2027T>A), and Southwest Asians (c.716T>A) were all present in Mongolian patients, demonstrating a panethnic mutation spectrum. Digenic inheritance was not observed. There was no clear genotype-phenotype correlation between specific *SLC26A4* genotypes and hearing levels or inner ear malformations.

**Conclusions:** This study provides a comprehensive overview of the genetic landscape of EVA in the Mongolian population. The identification of biallelic *SLC26A4* pathogenic variants in all families highlights the importance of this gene in the pathogenesis of EVA. The unique mutation spectrum observed in this study may reflect the genetic diversity resulting from historical migrations of Mongolians.

## Introduction

Sensorineural hearing loss (SNHL) affects an estimated 34 million children globally [1]. Approximately half of these cases are attributed to genetic factors [2, 3]. Recessive variants in the *SLC26A4* gene are the second most common cause of genetic hearing loss worldwide after *GJB2* [4]. This gene, located on chromosome 7 [5], encodes the pendrin protein, which is responsible for anion exchange in the inner ear [6-8], thyroid, and kidneys [5, 9]. Pathogenic variants in *SLC26A4* can cause autosomal recessive non-syndromic SNHL, DFNB4 (OMIM #600791) [10, 11] or Pendred syndrome (PS, OMIM #605646) [12-14]. DFNB4 is characterized by inner ear malformations, including enlarged vestibular aqueduct (EVA) and cochlear incomplete partition type II (IP-II). PS presents with similar inner ear malformations, but also includes thyroid goiter with iodine organification defect [11, 15].

To date, approximately 600 pathogenic or likely pathogenic variants in the *SLC26A4* gene have been documented in the variant databases ClinVar [16] and DVD [17] (last accessed on January 25, 2025). Different ethnic populations harbor different *SLC26A4* founder variants and mutation spectra [18]. For example, c.1246A>C (p.T416P) and c.1001G>A (p.G334E) are the predominant pathogenic variants in Caucasian populations, c.2168A>G (p.H723R) is prevalent in Japanese and Korean populations, whereas c.919-2A>G is most common in Han Chinese and Taiwanese populations [19-23].

In our previous study in 2018, we applied hot spot mutation screening in a Mongolian cohort of 188 SNHL patients and identified a unique *GJB2* mutation spectrum in Mongolian patients, which was significantly different from other Asian and European populations [24]. However, the information on *SLC26A4* mutations in Mongolian patients was rather limited at that time due to the unavailability of comprehensive sequencing tools (e.g., next-generation sequencing) and the lack of sophisticated phenotypic characterization (e.g., imaging studies in the hearing impaired patients). Recent advances in sequencing technology and increased accessibility of temporal bone imaging have facilitated more comprehensive genetic testing and phenotypic characterization of SNHL in Mongolia. This study re-examines the genetic landscape of *SLC26A4* variants in a Mongolian cohort confirmed with EVA on imaging studies using whole-exome sequencing (WES) to provide a more detailed understanding of the mutational spectrum within this population.

## Materials and Methods

### Subjects

Nineteen Mongolian patients from 15 unrelated families diagnosed with EVA with/without cochlear IP-II, as confirmed by high-resolution computed tomography (HRCT) imaging of the temporal bone, were recruited between 2022 and 2024. All participants provided written informed consent for genetic testing before undergoing WES. This study was approved by the research ethics committees of the National Taiwan University Hospital (NTUH) (202007065RINB) and the EMJJ Otolaryngology Hospital of Mongolia (Medical Ethics Committee of the Ministry of Health, Mongolia; No: 23/065).

### Clinical examinations

Detailed clinical data were collected from the proband and affected members of each family, including a comprehensive family history, personal medical history, and physical examination. Audiological evaluations were performed using pure-tone audiograms or auditory brainstem response (ABR), depending on the age or neurological status. Hearing levels were determined by averaging the air conduction thresholds at 0.5, 1, 2, and 4 kHz from both ears. Hearing levels were then classified as mild [26-40 decibel hearing level (dBHL)], moderate (41-60 dBHL), severe (61-80 dBHL), or profound (>81 dBHL) [25]. HRCT of the temporal bone with 1 mm contiguous axial and coronal sections was performed to evaluate the structure of the inner ear [26].

### Genetic sequencing and analysis

Genomic DNA was extracted from dried blood spot samples from each participant. WES was performed using the NovaSeq 6000 platform (Illumina Inc., San Diego, CA, USA). Detailed procedures for sample preparation and WES data processing are described in our previous study [27]. Identification of causative variants was based on several factors: minor allele frequency (MAF) of less than 1% in the gnomAD population database [28], correlation with patient phenotypes and medical records, and evidence from disease databases such as ClinVar [16] and the Deafness Variation Database (DVD) (version 9) [17]. Pathogenicity was assessed using a variety of prediction tools, including CADD (version 1.4) for all variant types; Sorting Intolerant From Tolerant (SIFT) (version 2019) [29] and Polymorphism Phenotyping v2 (PolyPhen-2) [30] for missense variants; and SpliceAI (version 1.3) [31] for intronic variants. Pathogenicity classification was performed according to the American College of Medical Genetics and Genomics (ACMG) guidelines [32].

### Statistical Analysis

Continuous variables are presented as mean ± standard deviation and were compared using the Wilcoxon signed-rank test. Categorical variables are presented as numbers and percentages and were analyzed using Fisher’s exact test. Statistical significance was defined as a p-value < 0.05. All statistical analyses were performed using R version 4.4.1 (Race for Your Life).

### Results

A total of 19 patients with EVA alone (n = 4) or combined with IP-II (n = 15) were recruited. The mean age was 8.2 years old, ranging from 2 to 16 years old. The gender distribution was 8 males (42.1%) and 11 females (57.9%). Four children (21.1%) had EVA alone with normal cochlear partition, while 15 (78.9%) had IP-II additionally. All participants had bilateral SNHL, with five children (26.3 %) having severe SNHL and fourteen children (73.7 %) having profound SNHL.

All probands of the 15 unrelated Mongolian families were confirmed to carry bi-allelic *SLC26A4* pathogenic variants (**Table 2**). These variants were either homozygous or compound heterozygous. The most common variant was c.919-2A>G, found in 12 out of 30 alleles (40%). The second most common variant was c.2027T>A (p.L676Q), found in 7 out of 30 alleles (23.3%). Other variants such as c.1318A>T (p.K440X), c.281C>T (p.T94I), c.716T>A (p.V239D), c.1229C>T (p.T410M), c.1547dup (p.S517FfsX10), c.1975G>C (p.V659L), and c.2089+1G>A were also identified.

**Table 1.** Demographic data of the 19 Mongolian patients.

| Variables | EVA alone<br>($n = 4$) | EVA with IP-II<br>($n = 15$) | Total<br>N = 19 | p-value |
| --- | --- | --- | --- | --- |
| | Mean ± SD | Mean ± SD | Mean ± SD | |
| Age, year | 13.4 ± 6.0 | 7.5 ± 4.9 | 8.2 ± 5.1 | 0.414 |
| Hearing level,<br>dBHL | 87.6 ± 6.6 | 89.4 ± 14.9 | 89.0 ± 13.4 | 0.721 |
|  | **n (%)** | **n (%)** | **n (%)** |  |
| Sex |  |  |  |  |
| Male | 2 (50.0) | 6 (40.0) | 8 (42.1) | 0.941 |
| Female | 2 (50.0) | 9 (60.0) | 11 (57.9) |  |

**Table 2.** *SLC26A4* genotypes, hearing levels, and imaging features of the 19 Mongolian patients.

| IDs* | Age range (Yrs) | Allele 1 |  | Allele 2 |  | Hearing loss level | Inner ear malformation |
| --- | --- | --- | --- | --- | --- | --- | --- |
|  |  | Nucleotide change | Amino acid change | Nucleotide change | Amino acid change |  |  |
| Patient 1 | 16-20 | c.919-2A>G | - | c.919-2A>G | - | profound | EVA |
| Patient 1-II | 11-15 | c.919-2A>G | - | c.919-2A>G | - | profound | EVA |
| Patient 2 | 11-15 | c.919-2A>G | - | c.281C>T | p.T94I | severe | EVA |
| Patient 3 | 1-5 | c.919-2A>G | - | c.2027T>A | p.L676Q | profound | EVA |
| Patient 4-I | 6-10 | c.1318A>T | p.K440X | c.1229C>T | p.T410M | profound | EVA & IP-II |
| Patient 4-II | 6-10 | c.1318A>T | p.K440X | c.1229C>T | p.T410M | profound | EVA & IP-II |
| Patient 5 | 1-5 | c.919-2A>G | - | c.716T>A | p.V239D | profound | EVA & IP-II |
| Patient 6 | 1-5 | c.2027T>A | p.L676Q | c.2027T>A | p.L676Q | profound | EVA & IP-II |
| Patient 7 | 1-5 | c.919-2A>G | - | c.2027T>A | p.L676Q | severe | EVA & IP-II |
| Patient 8 | 1-5 | c.919-2A>G | - | c.1547dup | p.S517FfsX10 | severe | EVA & IP-II |
| Patient 9 | 1-5 | c.919-2A>G | - | c.1318A>T | p.K440X | profound | EVA & IP-II |
| Patient 10 | 11-15 | c.919-2A>G | - | c.2027T>A | p.L676Q | profound | EVA & IP-II |
| Patient 11 | 11-15 | c.919-2A>G | - | c.919-2A>G | - | severe | EVA & IP-II |
| Patient 12 | 6-10 | c.1975G>C | p.V659L | c.2027T>A | p.L676Q | profound | EVA & IP-II |
| Patient 13-I | 11-15 | c.1318A>T | p.K440X | c.1318A>T | p.K440X | profound | EVA & IP-II |
| Patient 13-II | 16-20 | c.1318A>T | p.K440X | c.1318A>T | p.K440X | profound | EVA & IP-II |
| Patient 13-III | 6-10 | c.1318A>T | p.K440X | c.1318A>T | p.K440X | severe | EVA & IP-II |
| Patient 14 | 6-10 | c.2027T>A | - | c.2089+1G>A | - | profound | EVA & IP-II |
| Patient 15 | 1-5 | c.919-2A>G | p.L676Q | c.1318A>T | p.K440X | profound | EVA & IP-II |

EVA, enlarged vestibular aqueduct; IP-II, cochlear incomplete partition type II
\* All IDs are not represented as the hospital identities.

Among the 19 patients, 7 segregated homozygous variants: three individuals (patients 1-I, 1-2, and 11) with homozygous c.919-2A>G; three individuals (patient 13-I, 13-II, and 13-III) with homozygous c.1318A>T; and one individual (patient 6) with homozygous c.2027T>A. The remaining 12 patients segregated compound heterozygous *SLC26A4* variants. There is no clear correlation between specific *SLC26A4* genotypes and the level of hearing loss. Individuals with the same genotype, such as c.919-2A>G homozygotes, may have either profound or severe hearing loss. Notably, two boys aged in 11-15 years (Patient 11, *SLC26A4*:c.[919-2A>G];[919-2A>G]) and 6-10 years (Patient 12, *SLC26A4*:c.[919-2A>G];[2027T>A]) were born with normal hearing that subsequently deteriorated. This suggests that other factors, in addition to the *SLC26A4* genotype, may contribute to the variability in hearing loss levels.

Similarly, there is no clear correlation between specific *SLC26A4* genotypes and inner ear malformations. While some genotypes are exclusively associated with EVA alone, the small sample size and the presence of overlapping genotypes limit definitive conclusions.

## Discussion

In this study, we investigated the genetic basis of hearing loss in Mongolian patients with EVA with/without IP-II. We performed WES on 15 unrelated families and identified bi-allelic *SLC26A4* pathogenic variants in all of them (100%). The most prevalent variant was c.919-2A>G (40%), followed by c.2027T>A (23.3%) and c.1318A>T (16.7%).

The mutation spectrum of *SLC26A4* in our cohort showed unique characteristics compared to other populations. The c.919-2A>G variant, which is prevalent in East Asians, was also the most common in our study. In addition, we found the c.2027T>A variant, which is common in North Asians such as Tuvinian and Altai patients [33], and the rare c.716T>A variant, which was previously reported in Iranians [13, 34, 35]. This pan-ethnic spectrum may reflect the genetic diversity resulting from the historical migrations of the Mongols. Notably, the c.2168A>G variant, which is common in Japanese [21] and Korean [22] populations, was not detected in our cohort.

Our study achieved a high diagnostic yield (100%) for bi-allelic *SLC26A4* variants in Mongolian patients with EVA. Previous studies by Tsukamoto et al. [21] and Park et al. [22] showed that approximately 80% of EVA patients in Japanese and Korean populations were identified with bi-allelic pathogenic *SLC26A4* variants, 10% with mono-allelic pathogenic variants, and 10% with no detected variants. Similar studies conducted in European and North American populations showed that 25% of EVA patients had bi-allelic pathogenic *SLC26A4* variants, 25% had mono-allelic variants, and the remaining 50% had no detected pathogenic variants [36, 37]. Taken together, the diagnostic yield in our study is slightly higher than in other East Asian series and significantly higher than in the Caucasian series. In addition to the ethnic differences, the high diagnostic yield in our study could be attributed to the use of WES, which allows the detection of a wider range of variants compared to conventional hotspot sequencing.

Digenic inheritance, in which a mono-allelic *SLC26A4* variant is combined with another variant in genes such as *FOXI1* [38], *KCNJ10* [39], or *EPHA2* [40], has been proposed as a possible cause of DFNB4 and PS. However, we did not find any cases of digenic inheritance in our cohort. All patients with confirmed variants had bi-allelic *SLC26A4* mutations, and no pathogenic or likely pathogenic variants in *FOXI1, KCNJ10*, or *EPHA2* were detected. This suggests that digenic inheritance may play a less important role in the Mongolian population compared to other populations.

In this study, we did not observe clear correlations between specific *SLC26A4* genotypes and hearing features or inner ear malformations. Previous literature has investigated the association between specific *SLC26A4* genotypes and phenotypic variability, including the presence of Pendred syndrome and the severity of hearing loss. It has been reported that Pendred syndrome is more likely to be associated with biallelic *SLC26A4* variants than DFNB4 [15]. Certain *SLC26A4* genotypes have also been associated with less severe hearing loss [41, 42]. However, most other studies have not found a clear genotype-phenotype correlation [43-45]. This may be due to the allelic heterogeneity of *SLC26A4*, as many different *SLC26A4* variants have been identified and their effects on protein function may vary. In addition, environmental factors such as head trauma or exposure to loud noise may also contribute to hearing loss in individuals with *SLC26A4* variants [46, 47].

To our knowledge, this study is the first to comprehensively investigate the genetic landscape of EVA in the Mongolian population. Our use of WES allowed the detection of both common and rare variants, providing valuable insights into the genetic diversity of this population. The identification of a unique pan-ethnic mutational spectrum, including the c.2027T>A variant commonly found in North Asian populations and the c.716T>A variant previously reported in Southwest Asian populations, highlights the potential influence of historical migrations on the genetic makeup of Mongolians. However, our study is limited by its small sample size, which may prevent the detection of additional variants contributing to EVA in this population, as well as the delineation of genotype-phenotype correlations. Further studies with larger cohorts are needed to fully elucidate the genetic complexity of hereditary hearing impairment in Mongolians.

## Conclusion

This study provides a comprehensive overview of the genetic landscape of EVA in the Mongolian population. The identification of biallelic *SLC26A4* pathogenic variants in all families highlights the importance of this gene in the pathogenesis of EVA. The unique mutation spectrum observed in this study may reflect the genetic diversity resulting from historical migrations of Mongolians. Further studies with larger cohorts are needed to fully elucidate the genetic complexity of EVA in this population.

## Data Availability

All data produced in the present study are available upon reasonable request to the authors.

## Ethical approval and consent to participate

The study was approved by the NTUH Research Ethics Committee (202007065RINB) and EMJJ Otolaryngology Hospital of Mongolia (Medical Ethics Committee of the Ministry of Health, Mongolia (No: 23/065)). Informed consent was obtained from all participants and/or their legal guardians.

## Availability of data and materials

The datasets used and/or analyzed during the current study are available from the corresponding authors on reasonable request.

## Authors’ contributions (CRediT author statement)

- Conceptualization: *Jargalkhuu Erdenechuluun*, *Bayasgalan Gombojav*
- Methodology: *Bayasgalan Gombojav, Cheng-Yu Tsai, Chien-Hsing Lin*
- Software: *Bayasgalan Gombojav*
- Validation: *Yue-Sheng Lu, Ho-Peng Hsu*
- Formal analysis: *Jargalkhuu Erdenechuluun, Bayasgalan Gombojav*, *Tserendulam Batsaikhan, Zaya Makhbal, Narandalai Danshiitsoodol, Maralgoo*,*Jargalmaa, Tuvshinbayar Jargalkhuu*
- Investigation: *Tserendulam Batsaikhan, Cheng-Yu Tsai*
- Resources: *Pei-Hsuan Lin, Hung-Ju Su, Chuan-Jen Hsu, Pei-Lung Chen*
- Data Curation: *Jargalkhuu Erdenechuluun, Bayasgalan Gombojav, Tserendulam Batsaikhan*
- Writing (Original Draft): *Jargalkhuu Erdenechuluun, Bayasgalan Gombojav, Tserendulam Batsaikhan*
- Writing (Review/Editing): *Bayasgalan Gombojav, Tserendulam Batsaikhan, Cheng-Yu Tsai, Chen-Chi Wu*
- Visualization: *Jargalkhuu Erdenechuluun, Bayasgalan Gombojav*
- Supervision: *Jargalkhuu Erdenechuluun, Chen-Chi Wu*
- Project administration: *Cheng-Yu Tsai, Yue-Sheng Lu, Ho-Peng Hsu, Tserendulam Batsaikhan Zaya Makhbal, Maralgoo*,*Jargalmaa*
- Funding acquisition: *Chen-Chi Wu, Jargalkhuu Erdenechuluun*

## Acknowledgements

We sincerely thank the A1 Laboratory of Genetic Testing at the National Taiwan University Hospital and Phalanx Biotech Inc. for their invaluable experimental and technical support. We would also like to thank all the participating subjects and their families for their generous contributions to this study.

